# How to mitigate selection bias in COVID-19 surveys: evidence from five national cohorts

**DOI:** 10.1101/2024.03.06.24303781

**Authors:** Martina K. Narayanan, Brian Dodgeon, Michail Katsoulis, George B. Ploubidis, Richard J. Silverwood

**Affiliations:** Centre for Longitudinal Studies, Institute of Education, University College London; MRC Unit for Lifelong Health & Ageing

**Keywords:** Cohort studies, COVID-19, Longitudinal data, Non-response, Missing data, Multiple imputation, Weighting

## Abstract

**Background:** Non-response is a common problem, and even more so during the COVID-19 pandemic where social distancing measures challenged data collections. As non-response is often systematic, meaning that respondents are usually healthier and from a better socioeconomic background, this potentially introduces serious bias in research findings based on COVID-19 survey data. The goal of the current study was to see if we can reduce bias and restore sample representativeness despite systematic non-response in the COVID-19 surveys embedded within five UK cohort studies using the rich data available from previous time points.

**Methods:** A series of three surveys was conducted during the pandemic across five UK cohorts: National Survey of Health and Development (NSHD, born 1946), 1958 National Child Development Study (NCDS), 1970 British Cohort Study (BCS70), Next Steps (born 1989-90) and Millennium Cohort Study (MCS, born 2000-02). We applied non-response weights and utilised multiple imputation, making use of covariates from previous waves which have been commonly identified as predictors of non-response, to attempt to reduce bias and restore sample representativeness.

**Results:** Response rates in the COVID-19 surveys were lower compared to previous cohort waves, especially in the younger cohorts. We identified bias due to systematic non-response in the distributions of variables including parental social class and childhood cognitive ability. In each cohort, respondents of the COVID-19 survey had a higher percentage of parents in the most advantaged social class, and a higher mean of childhood cognitive ability, compared to the original (full) cohort sample. The application of non-response weights and multiple imputation was successful in reducing bias in parental social class and childhood cognitive ability, nearly eliminating it for the former.

**Conclusions:** The current paper demonstrates that it is possible to reduce bias from non-response and to a large degree restore sample representativeness in multiple waves of a COVID-19 survey embedded within long running longitudinal cohort studies through application of non-response weights or multiple imputation. Such embedded COVID-19 surveys therefore have an advantage over cross-sectional COVID-19 surveys, where non-response bias cannot be handled by leveraging previously observed information on non-respondents. Our findings suggest that, if non-response is appropriately handled, analyses based on the COVID-19 surveys within these five cohorts can contribute significantly to COVID-19 research, including studying the medium and long-term effects of the pandemic.

## 1. Introduction

Since the emergence of COVID-19, an increasing amount of research has studied the impact of the pandemic. Most of that research is based on web surveys, phone surveys and other selective samples which have been recruited for the first time during the pandemic ^1,2^. Commonly, those selective samples may not be representative of the entire population, since respondents are often systematically different from non-respondents ^3^. Such non-representativeness has the potential to introduce bias into analyses undertaken in these datasets. It is difficult to sufficiently address the bias caused by selective response when there is no information on the people who chose not to respond. Approaches for doing so often rely on reweighting the achieved sample to be representative of a given population in terms of the overall distributions of certain variables, for example the proportion of individuals living in different geographical areas or the proportion of people working in different sectors. However, the availability of such population information may be limited, especially during the pandemic, which has affected the response in benchmark population surveys as well ^4^. Therefore, the reweighted sample may be unable to fully capture the nuances of the population, meaning that bias due to selection remains. COVID-19 surveys conducted within the context of pre-existing longitudinal population-based surveys provide an alternative approach which potentially allows us to capitalise on the rich data available in earlier waves to correct for bias in the embedded COVID-19 survey.

We consider five UK longitudinal cohort studies – the National Survey of Health and Development (NSHD) ^5,6^, the 1958 National Child Development Study (NCDS) ^7^, the 1970 British Cohort Study (BCS70) ^8,9^, Next Steps ^10^, and the Millennium Cohort Study (MCS) ^11,12^ – with participants aged between 19 (MCS) and 74 (NSHD) at the start of the COVID-19 pandemic. Three COVID-19-specific surveys, between May 2020 and March 2021, collected insights into the lives of the participants of all five cohort studies during the pandemic, including their physical and mental health and wellbeing, family and relationships, education, work, and finances ^13^. As is common in such surveys, particularly during the pandemic, there was substantial non-response which requires appropriate handling^4^. Missing values due to non-response mean less efficient estimates because of the reduced size of the analysis sample, but also introduce the potential for bias. Two common approaches for handling non-response include weighting ^14,15^ and multiple imputation (MI) ^16,17,18^.

In this paper we aim to describe the response to the COVID-19 surveys across the five cohorts and detail the implementation of non-response weights and MI to handle missing data, capitalising on the rich data cohort members provided over the years prior to the COVID-19 surveys in order to restore sample representativeness. Showing that sample representativeness can be restored for these COVID-19 surveys is especially important for future research studying the medium and long-term effects of the pandemic. This work builds upon recent work on appropriately handling non-response in NCDS, Next Steps and BCS70 ^19,20,21^.

## 2. Methods

### 2.1 Data

We use information from five nationally representative cohort studies, whose participants have been providing information about their lives since childhood. Brief details of the studies are given here; full details are available elsewhere ^5–8,11,12^.

#### 2.1.1 National Survey of Health and Development (NSHD)

The NSHD is a representative sample (N=5362) of men and women born in England, Scotland, and Wales in March 1946 ^5,6^. Data were collected from birth and study members have been followed up 24 times. At the first wave of the COVID-19 survey cohort members were around 73 years old.

#### 2.1.2 1958 National Child Development Study (NCDS)

The NCDS is a representative sample of 17,500 babies born in England, Scotland, and Wales in one week of 1958 ^7^. The birth survey has been followed by ten further data collections. At the first wave of the COVID-19 survey cohort members were around 62 years old.

#### 2.1.3 1970 British Cohort Study (BCS70)

The BCS70 follows the lives of more than 17,000 people born in England, Scotland, and Wales in a single week of 1970 ^8^. Following the birth survey there have so far been eight more surveys. At the first wave of the COVID-19 survey cohort members were around 50 years old.

#### 2.1.4 Next Steps

Next Steps, previously known as the Longitudinal Study of Young People in England (LSYPE), follows the lives of around 16,000 people in England born in 1989-90 ^10^. Next Steps cohort members have been surveyed 8 times starting at 14 years old. At the first wave of the COVID-19 survey cohort members were around 31 years old.

#### 2.1.5 Millennium Cohort Study (MCS)

The MCS is following the lives of around 19,000 young people born across England, Scotland, Wales, and Northern Ireland in 2000-02 ^11,12^. The first data collection took place at 9 months with six follow up surveys since then. At the first wave of the COVID-19 survey cohort members were around 20 years old.

#### 2.1.6 COVID-19 surveys

A series of three surveys was run across all five cohorts during the pandemic ^13^. A first COVID-19 survey (Wave 1) took place in May 2020 at the time when the UK was in a first national lockdown, with over 15,000 study participants taking part across the five cohorts. Nearly 20,000 participants took part in a second survey (Wave 2) in September/October 2020 during a period in which lockdown restrictions had been mostly lifted. The Wave 3 survey took place in February/March 2021, during the third UK lockdown, with over 22,000 participants. The Wave 1 and Wave 2 surveys were conducted purely online. In Wave 3 participants were initially invited to take part online but a subset of web-survey non-respondents were subsequently followed up and invited to take part via telephone.

The Wave 1 survey invitations were sent via email, meaning the issued sample comprised all cohort members for whom an email address was held, provided that they a) had not permanently withdrawn from the study, b) were not ‘permanently untraced’ and c) were not known to have died. At Waves 2 and 3 it was possible to include those for whom no email address was held through use of postal invitations and cohort members with no email address were invited to take part provided that they had taken part in a recent major sweep of data collection or their address had been recently confirmed. At Waves 2 and 3 study members who had ‘opted out’ of the COVID-19 project in previous waves were not invited to take part. A t Wave 3, for NSHD only, the issued sample was restricted to individuals who had responded to the COVID-19 Wave 1 and/or 2 surveys.

Emigrants for whom an email address was held were included in the issued sample. This includes study members living outside of Great Britain in the case of NCDS, BCS70 and Next Steps and those living outside the UK (i.e. including Northern Ireland) in the case of MCS. MCS parents were also invited to complete the surveys but are not considered further in the present study.

For the purpose of weighting and MI, the target population of each cohort is identified as individuals born in the specified birth period of the cohort who are alive and still residing in the UK. Information on mortality and emigration was not available for MCS and Next Steps, and we therefore did not adjust the target populations to take deaths or emigrations into account. We expect mortality in both cohorts to be very low, and rates of emigration are also unlikely to be very significant.

However, to the extent that the target population in MCS and Next Steps may have been overestimated due to these factors, this would lead to a (likely, minor) underestimation of response relative to target in these cohorts. In MCS there was an additional exclusion from the target population: only singletons and one twin or triplet from each twin pair/triplet set were included (i.e. second twin and second/third triplets were excluded). Further details of the three COVID-19 surveys can be found elsewhere ^13^.

### 2.2 Measures

#### 2.2.1 Covariates

A list of covariates included in the derivation of non-response weights and in imputation models for the restoring representativeness examples can be found in Table 1. More detailed information about the coding of covariates is included in Table S1 (Supplementary Material).

**Table 1.** Variables included in the weight derivation models and imputation models.

|  | NSHD | NCDS | BCS70 | Next Steps | MCS |
| --- | --- | --- | --- | --- | --- |
| Sex | Birth | Birth | Birth | Age 14 | 9 months |
| Ethnicity | - | - | - | Age 14 | 9 months<br>Age 3 |
| Parental social class | Age 4<sup>G</sup><br>Age 11<sup>F</sup> | Birth<br>Age 11<sup>F</sup> | Birth<br>Age 10<sup>F</sup> | Age 14<sup>F</sup> | 9 months<br>Age 11<sup>F</sup> |
| Number of rooms at home/persons per room | Birth | Birth | Birth | - | 9 months |
| Cognitive ability | Age 8<sup>F</sup><br>Age 11 | Age 7<sup>F</sup><br>Age 11 | Age 10<br>Age 5<sup>F</sup> | - | Age 5<sup>F</sup><br>Age 7 |
| Early life mental health | Age 13 & 15 | Age 16 | Age 16 | Age 15 | Age 11 |
| Voting | Age 26 | Age 42 | Age 42 | Age 20 | NA |
| Membership in organisations | Age 43 | Age 42 | Age 42 | Age 26 | Age 14 |
| Internet access prior to web survey | Age 69 | Age 50 | Age 46 | Age 26 | Age 14 |
| Consent for biomarkers | Age 60-64<sup>B</sup> | Age 44 | Age 46 | - | - |
| Consent for linkages | Age 60-64<sup>B</sup> | - | - | Age 26 | - |
| Educational qualifications | Age 26 | Age 42 | Age 42 | Age 26 | 9 months<sup>A</sup> |
| Economic activity | Age 60-64 | Age 50 | Age 46 | Age 26 | Age 14<sup>A</sup> |
| Partnership status | Age 69 | Age 50 | Age 46 | Age 26 | Age 14 |
| Psychological distress | Age 69 | Age 50 | Age 46 | Age 26 | Age 14 |
| BMI | Age 69 | Age 50 | Age 46 | Age 26 | Age 11 |
| Self-rated health | Age 69 | Age 50 | Age 46 | Age 26 | Age 14 |
| Smoking status | Age 69 | Age 50 | Age 46 | Age 26 | Age 14 |
| Maternal mental health<sup>C</sup> | - | - | - | - | 9 months |
| Social capital/social support | Age 69 | Age 50 | Age 46 | Age 26 | Age 14 |
| Income | Age 69 | Age 55 | Age 42 | Age 26 | Age 14<sup>A</sup> |
| Number of non-responses across all previous sweeps | Birth – age 69 | Birth – age 55 | Birth – age 42 | Age 14 – age 26 | 9 months – age 14 |
| Response to COVID-19 Wave 1 survey<sup>D</sup> | Age 74 | Age 62 | Age 50 | Age 30 | Age 19 |
| Response to COVID-19 Wave 2 survey<sup>E</sup> | - | Age 62 | Age 50 | Age 30 | Age 19 |

<sup>A</sup> Main respondent, >90% mothers.
<sup>B</sup> Excluded from final model due to collinearity.
<sup>C</sup> Also available in BCS70 at age 16 but not included in model.
<sup>D</sup> Included in Wave 2 and 3 response models only.
<sup>E</sup> Included in Wave 3 response model only, apart from in NSHD where Wave 3 web survey was only issued to those who had responded to previous COVID-19 surveys.
<sup>F</sup> These were used as variables in the restoring sample representativeness examples, which means they were not included in the derivation of weights.
<sup>G</sup> Not included in multiple imputation model due to convergence issues.
NSHD: National Survey of Health and Development; NCDS: 1958 National Child Development Study; BCS70: 1970 British Cohort Study; MCS: Millennium Cohort Study.

#### 2.2.2 Parental social class in childhood

The true distribution of parental social class in each cohort sample is known, as the variable is observed in childhood in nearly all participants in each cohort. This serves as a comparator to examine potential bias in the distribution of parental social class among respondents to the COVID-19 surveys, allowing us to examine how non-response weighting and MI can help correct that bias.

For these analyses, we chose parental social class variables in childhood rather than at birth, as the first wave of Next Steps was at age 14. In NSHD, parental social class was measured as father’s social class at age 11 and coded in three categories (professional/intermediate, skilled, and partly-/unskilled). For NCDS, it was father’s or male head of household’s social class at age 11, coded in the same three categories. For BCS70, it was father’s (or mother’s if father’s information was missing) social class at age 10, again coded in the same three categories. For Next Steps, we measured father’s social class at age 14, which was coded in four categories (managerial, intermediate, routine/semi-routine, and never worked). For MCS, it was the highest parental social class at age 11, coded in three categories (managerial, intermediate, and routine/semi-routine). As all analyses in the current paper were run separately for each cohort, we did not attempt to further harmonise the variables for parental social class.

#### 2.2.3 Childhood cognitive ability

Similar to parental social class, we wanted to show how non-response weighting and MI can restore representativeness for childhood cognitive ability given selective response to the COVID-19 surveys. In NSHD, cognitive ability was measured as a standardised score at age 8 based on four different tests (Reading Comprehension, Word Reading, Vocabulary and Picture Intelligence) ^22^. For NCDS, we used a standardised score at age 7 based on a different set of four tests (Southgate Group Reading Test, Copying Designs Test, Human Figure Drawing, Problem Arithmetic Test) ^23^. For BCS70, we used a standardised score at age 5 based on 3 tests (English Picture Vocabulary Test, Copying Designs Test, Human Figure Drawing) ^24^. For MCS, we used a standardised score at age 5 based on three tests (BAS II Naming Vocabulary, BAS II Pattern Construction, BAS II Picture Similarities) ^25^. Next Steps does not have any measures of childhood cognitive ability available and was therefore not included in these analyses. Again, as all analyses in the current paper were run separately for each cohort, we did not attempt to further harmonise the variables.

### 2.3 Statistical methods

#### 2.3.1 Derivation of non-response weights

The derivation of the COVID-19 survey non-response weights was implemented in each cohort separately but following a common approach. For each wave separately, we proceeded as follows: 1) Within the sample corresponding to the target population (those alive and living in the UK), we modelled COVID-19 survey response conditional on a common set of covariates using logistic regression. The selection of covariates was informed by results of the CLS Missing Data Strategy ^19,20,21^ and their *a priori* assumed association with the probability of response and/or with key COVID-19 survey variables. 2) For COVID-19 survey respondents, we predicted the probability of response from the model. 3) We then calculated the COVID-19 survey non-response weight as the inverse of the probability of response. 4) We examined the distribution of derived non-response weights across cohorts to decide whether truncation may be desirable, applying this if so. 5) Finally, we calibrated the COVID-19 survey non-response weights so that they summed to the number of COVID-19 survey respondents in each cohort.

We aimed to use broadly the same set of variables in each cohort to ensure consistency in the non-response weight derivation (see Table 1). However, it was not possible to include identical sets of variables due to data being collected at different ages and using different questions, and occasionally due to certain variables not been collected at all in some cohorts. Given that the non-response weight derivation was implemented separately in each cohort, such relatively minor differences were not deemed likely to be important.

When deriving the non-response weights, missing values in the covariates were handled using MI, conducted in each cohort separately. The imputation model for each cohort included all the covariates, response at the wave under consideration and, for relevant cohorts (NSHD, Next Steps and MCS), the design weight. Five imputed datasets were created using chained equations. Such a small number of imputations was deemed sufficient as only point estimates (the probability of COVID-19 survey response) were to be estimated from the MI analysis (i.e. no inferences were being made). Models for COVID-19 survey response were fitted in each imputed dataset and combined using standard rules. For further details of the derivation of weights, including estimated response models and distributions of non-response weights prior to and post truncation, see the COVID-19 Survey User Guide ^13^.

The purpose of this paper is to show that sample representativeness can be restored in the COVID-19 survey waves for parental social class and childhood cognitive ability when using non-response weights or MI. In some cases, the original non-response weights as documented in the COVID-19 Survey User Guide ^13^ included the exact same measures of parental social class or childhood cognitive ability in their derivation models that are also used as examples in the current paper (parental social class in Next Steps and MCS and childhood cognitive ability in NSHD, NCDS and MCS). For these specific cases, we created new non-response weights based on models which did not include the particular variable of interest in their derivation models. Wherever available, we included a similar variable from a different wave instead (e.g. for our example to restore sample representativeness for cognitive ability in NCDS at age 7, we did not include cognitive measures from age 7 but instead from age 11). For more details on the included covariates see Table 1. We conducted sensitivity analyses providing estimates based on the original and the newly created non-response weights; for full details see Methods S1 (Supplementary Material).

#### 2.3.2 Multiple imputation (MI)

In parallel analyses, we also utilised MI to restore sample representativeness of parental social class and childhood cognitive ability. MI was conducted separately for each cohort, with each imputation model containing the analysis variable of interest (parental social class in childhood or childhood cognitive ability) as well as all covariates used in the derivation of the non-response weights as described above. That way, non-response weights and MI models were based on the same set of variables, allowing meaningful comparison (see Table 1). MI was conducted using chained equations and 50 imputed datasets, and missing values were imputed for all individuals in the COVID-19 survey target population for each study (as defined above) to again ensure comparability across methods. Where applicable, design weights were included in the imputation model (for NSHD, Next Steps and MCS) to account for the survey structure.

#### 2.3.3 Restoring sample representativeness of childhood social class and cognitive ability

We conducted a series of analyses to examine the effectiveness of the derived non-response weights and the MI procedure in restoring sample representativeness for the distribution of parental social class in childhood and childhood cognitive ability. For each wave of the COVID-19 survey separately, we compared the distribution of parental social class across all cohort members to the distribution of the same variable in COVID-19 survey respondents only (to assess the extent of bias caused by non-response) and in COVID-19 survey respondents after the application of the non-response weights or MI procedure (to assess to what extent the bias due to non-response could be overcome). We conducted similar analyses for childhood cognitive ability.

All analyses were run using Stata SE 18.0 ^26^.

## 3. Results

### 3.1 COVID-19 survey response

The number of cohort members in the Wave 1, 2 and 3 target populations and the number of responses both within the issued samples and within the target populations are presented in Table 2. The total response rates of all cohort members with respect to the issued sample increased over time (37.5% in Wave 1, 39.1% in Wave 2 and 43.8% in Wave 3) and was strongly patterned by cohort (or, equivalently, by age) within each wave (e.g. 68.3% for NSHD through to 26.6% for MCS in Wave 1). The total response rates of all cohort members with respect to the target population (20.8% in Wave 1, 27.7% in Wave 2 and 31.2% in Wave 3) were markedly lower than those with respect to the issued sample due to cohort members within the target population not being within the issued sample for one of the reasons described above.

**Table 2.**
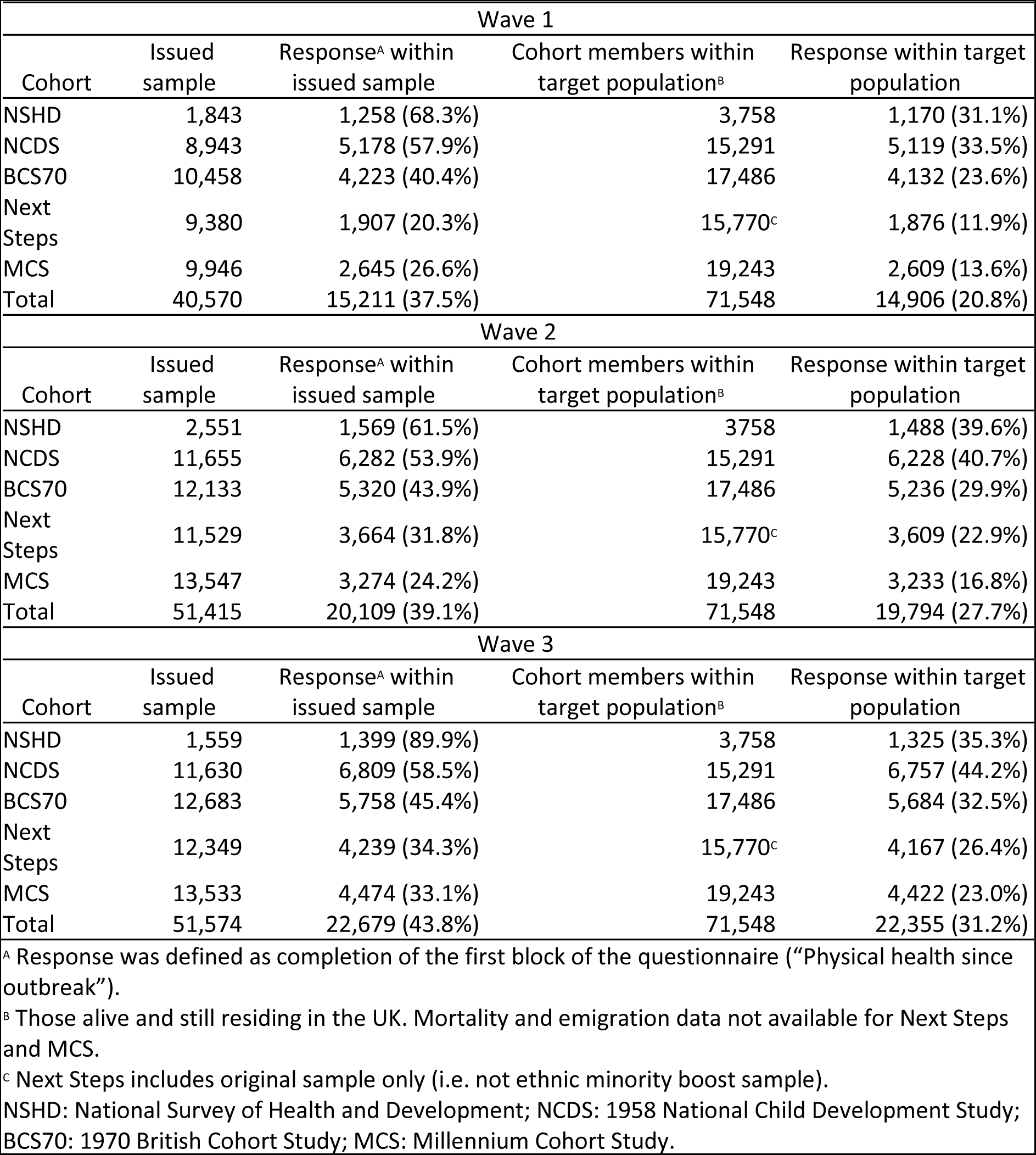
COVID-19 Wave 1, 2 and 3 surveys: issued sample, target population and response by cohort.

| Wave 1 |  |  |  |  |
| --- | --- | --- | --- | --- |
| Cohort | Issued sample | Response<sup>A</sup> within issued sample | Cohort members within target population<sup>B</sup> | Response within target population |
| NSHD | 1,843 | 1,258 (68.3%) | 3,758 | 1,170 (31.1%) |
| NCDS | 8,943 | 5,178 (57.9%) | 15,291 | 5,119 (33.5%) |
| BCS70 | 10,458 | 4,223 (40.4%) | 17,486 | 4,132 (23.6%) |
| Next Steps | 9,380 | 1,907 (20.3%) | 15,770<sup>C</sup> | 1,876 (11.9%) |
| MCS | 9,946 | 2,645 (26.6%) | 19,243 | 2,609 (13.6%) |
| Total | 40,570 | 15,211 (37.5%) | 71,548 | 14,906 (20.8%) |
| Wave 2 |  |  |  |  |
| Cohort | Issued sample | Response<sup>A</sup> within issued sample | Cohort members within target population<sup>B</sup> | Response within target population |
| NSHD | 2,551 | 1,569 (61.5%) | 3,758 | 1,488 (39.6%) |
| NCDS | 11,655 | 6,282 (53.9%) | 15,291 | 6,228 (40.7%) |
| BCS70 | 12,133 | 5,320 (43.9%) | 17,486 | 5,236 (29.9%) |
| Next Steps | 11,529 | 3,664 (31.8%) | 15,770<sup>C</sup> | 3,609 (22.9%) |
| MCS | 13,547 | 3,274 (24.2%) | 19,243 | 3,233 (16.8%) |
| Total | 51,415 | 20,109 (39.1%) | 71,548 | 19,794 (27.7%) |
| Wave 3 |  |  |  |  |
| Cohort | Issued sample | Response<sup>A</sup> within issued sample | Cohort members within target population<sup>B</sup> | Response within target population |
| NSHD | 1,559 | 1,399 (89.9%) | 3,758 | 1,325 (35.3%) |
| NCDS | 11,630 | 6,809 (58.5%) | 15,291 | 6,757 (44.2%) |
| BCS70 | 12,683 | 5,758 (45.4%) | 17,486 | 5,684 (32.5%) |
| Next Steps | 12,349 | 4,239 (34.3%) | 15,770<sup>C</sup> | 4,167 (26.4%) |
| MCS | 13,533 | 4,474 (33.1%) | 19,243 | 4,422 (23.0%) |
| Total | 51,574 | 22,679 (43.8%) | 71,548 | 22,355 (31.2%) |

<sup>A</sup> Response was defined as completion of the first block of the questionnaire (“Physical health since outbreak”).
<sup>B</sup> Those alive and still residing in the UK. Mortality and emigration data not available for Next Steps and MCS.
<sup>C</sup> Next Steps includes original sample only (i.e. not ethnic minority boost sample).
NSHD: National Survey of Health and Development; NCDS: 1958 National Child Development Study; BCS70: 1970 British Cohort Study; MCS: Millennium Cohort Study.

### 3.2. Restoring sample representativeness for parental social class

The results regarding parental social class in childhood are presented in Fig. 1 for Wave 1. Whilst the parental social class variables were not fully harmonised across cohorts, the percentage of cohort members in the highest social class (professional/managerial) was higher in the more recent (younger) cohorts. The extent of bias in the estimated percentage of cohort members in the highest social class caused by non-response to the COVID-19 Wave 1 survey varied across cohorts but was substantial in all cases. The bias was in the expected direction, with respondents to the COVID-19 Wave 1 survey disproportionately coming from a higher social class. However, the application of the non-response weights greatly reduced this bias, almost eliminating it in all cohorts so that the sample representativeness with respect to this variable was essentially restored. MI achieved very similar estimates to those using non-response weights (marginally closer to the known truth in some cohorts, marginally further away in others), also showing that sample representativeness can be restored successfully. Results for Wave 2 and 3 (Fig. S1 and S2, Supplementary Materials) were very similar.

**Fig. 1.**
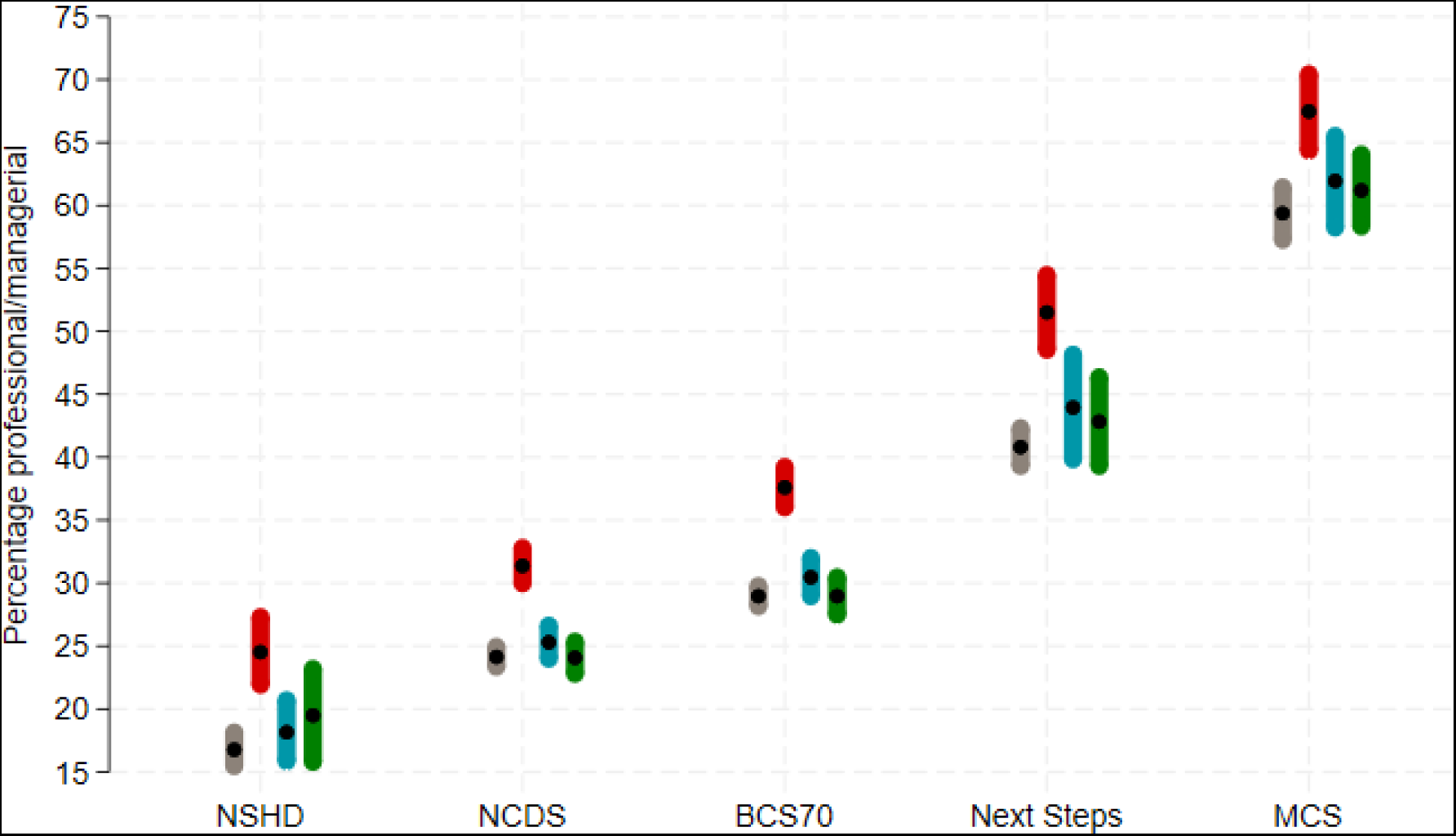
Percentage of highest social class (professional/managerial) in each cohort under different estimation approaches to account for non-response in the COVID-19 Wave 1 survey. Grey: using observed baseline data from the whole cohort; red: using observed baseline data from COVID-19 Wave 1 survey respondents only – unweighted (NCDS and BCS70) or using design weight only (NSHD, Next Steps and MCS); blue: using observed baseline data from COVID-19 Wave 1 survey respondents only – weighted using non-response weights (in addition to design weights as appropriate); green: using multiple imputation (plus design weight as appropriate). NSHD: National Survey of Health and Development; NCDS: 1958 National Child Development Study; BCS70: 1970 British Cohort Study; MCS: Millennium Cohort Study.

### 3.3. Restoring sample representativeness for childhood cognitive ability

The results regarding childhood cognitive ability at COVID-19 survey Wave 1 are presented in Fig. 2 Again, we see a substantial bias caused by non-response, with respondents showing a higher mean of childhood cognitive ability as compared to the original sample. Applying non-response weights and MI both greatly reduce the bias in all cohorts. While the bias is not fully removed for NCDS, BCS70 and MCS, MI estimates for NSHD show that the bias is near eliminated. Results for Wave 2 and 3 (Fig. S3 and S4, Supplementary Material) were very similar to Wave 1.

**Fig. 2.**
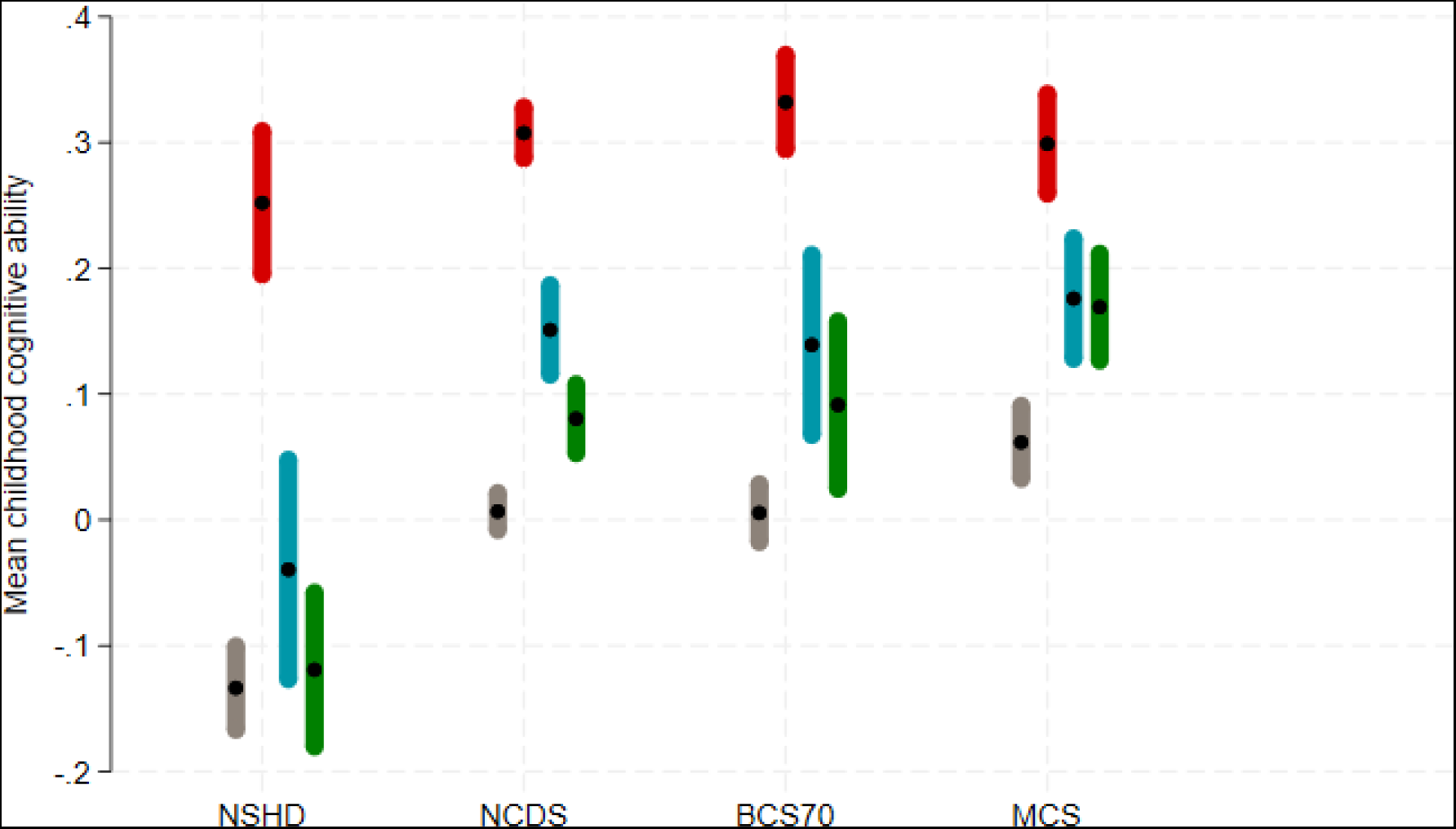
Mean of childhood cognitive ability in each cohort under different estimation approaches to account for non-response in the COVID-19 Wave 1 survey. Grey: using observed baseline data from the whole cohort; red: using observed baseline data from COVID-19 Wave 1 survey respondents only – unweighted (NCDS and BCS70) or using design weight only (NSHD, Next Steps and MCS); blue: using observed baseline data from COVID-19 Wave 1 survey respondents only – weighted using non-response weights (in addition to design weights as appropriate); green: using multiple imputation (plus design weight as appropriate). NSHD: National Survey of Health and Development; NCDS: 1958 National Child Development Study; BCS70: 1970 British Cohort Study; MCS: Millennium Cohort Study. Design weights were used in the estimation of means when available (NSHD, MCS) which explains why the mean of the standardised score is not always exactly 0.

### 3.4 Sensitivity analyses

Sensitivity analyses were run to compare results for newly created non-response weights with the original non-response weights from the COVID-19 Survey User Guide. For our childhood social class example (Fig. 5 Supplementary Material) the newly created non-response weights produced similar results as compared to the original non-response weights. For our childhood cognitive ability example (Fig. 6 Supplementary Material) newly created and original non-response weights produced similar results as well, as long as we included cognitive measures from other waves. For more details see Supplementary Materials.

## 4. Discussion

Although response rates in these COVID-19 surveys were comparable with other surveys carried out at the same time in the UK ^4^, they were lower compared to those in pre-pandemic waves of the same studies ^13^. We have shown that these relatively low response rates cause significant bias in the composition of the sample with respect to parental social class and childhood cognitive ability. Furthermore, we show that the application of non-response weights and MI can help significantly reduce, or even eliminate, non-response bias.

Response rates to the COVID-19 surveys varied across the COVID-19 waves, from 62% to 90% for NSHD, 54% to 59% for NCDS, 40% to 45% for BCS70, 20% to 34% for Next Steps, and 24% to 33% for MCS (all relative to the issued sample). Comparing that to response rates pre pandemic, with 84% for NSHD in 2014/16, 58% for NCDS in 2013/14, 70% for BCS70 in 2016, 49% for Next Steps in 2015, and 73% for MCS in 2018 ^13^, it appears that for BCS70, Next Steps and MCS (i.e. the younger cohorts) in particular, response was lower than what would be expected at a conventional sweep of data collection. Similarly, the Office for National Statistics (ONS) reports that for the Labour Force Survey, Survey of Living Conditions, Wealth and Assets Survey, and National Survey for Wales there was an increase in the proportion of respondents aged 46 years and over, and a decrease in the proportion of those aged 0 to 15 years and 16 to 45 years when data collections had to be adapted due to the start of the pandemic ^4^. It appears that especially for younger generations, data collected during the pandemic faced increased issues of non-response and thus an increased risk of bias in findings based on those data.

We did find bias due to non-response for our chosen examples, parental social class and childhood cognitive ability. In each cohort, respondents of the COVID-19 survey had a higher percentage of parents in the most advantaged social class, and a higher mean of childhood cognitive ability as compared to the original cohort sample. This is in agreement with the literature on non-response in longitudinal surveys in which those from a advantaged socio-economic background and higher cognitive ability are generally more likely to respond ^19,20,21^.

The application of non-response weights and MI was successful in reducing bias in parental social class and childhood cognitive ability, nearly eliminating it for the former. These serve as examples to show how the application of these approaches can reduce bias and increase sample representativeness for the COVID-19 survey waves. Our findings are in alignment with previous practical examples of the effectiveness of non-response weights and MI in the British cohort studies^19,20,21^.

In our analyses we used the same auxiliary variables in the imputation phase of our MI approach as were used in the previous derivation of the generic non-response weights in order to maintain comparability between the two methods. In practice, users of the data may choose to include more analysis-specific information in their non-response handling, either through inclusion of additional auxiliary variables in their MI analysis (which is an inherently analysis-specific approach) or by deriving their own analysis-specific weights, with further variables included in the response model. In many settings, such analysis-specific approaches may perform better than application of the generic non-response weights.

### Strengths and limitations

We were able to show that we can restore sample representativeness for COVID-19 surveys embedded within five ongoing UK cohorts using two different approaches to deal with missingness, making our findings more robust. Despite this the study has a few limitations. Firstly, we only looked at two specific childhood variables to see if we can restore sample representativeness, therefore we are cautious about generalising the results to other sample characteristics. Furthermore, our examples only included proportions and means of single variables, not more complex analytic estimates such as regression models including multiple variables. Secondly, although we did include design weights (for NSDH, Next Steps and MCS) in the MI models as well as the MI step of the weight derivation, we were not able to fully account for the survey structure in the imputation phase.

However, survey structure was correctly accounted for in the analytic phase. Lastly, instead of going back in time and looking at childhood variables, the current paper would have benefited from demonstrating sample representativeness in the COVID-19 surveys relative to external population benchmarks, such as from the Office of National Statistics (ONS). Unfortunately, data collections within ONS surveys were significantly affected during the pandemic as well, with interviews first pausing and eventually switching from in person to telephone interviews, which did result in less reliable population estimates during this period ^4^.

### Implications

The current paper demonstrates that it is possible to reduce bias from non-response and to a large degree restore sample representativeness in multiple waves of a COVID-19 survey embedded within ongoing longitudinal studies using the rich information collected at earlier waves through application of non-response weights or MI. Doing so, research findings based on the COVID-19 surveys within these five cohorts have a clear advantage relative to research based on samples collected during the pandemic only, and can contribute significantly to COVID-19 research by producing less biased research outputs. The findings have implications for future analyses exploring the medium and longer-term consequences of COVID-19 infection and the pandemic more broadly using data collected at subsequent waves.

## Data Availability

Data from NCDS, BCS, Next Steps and MCS are available through the UK Data Service (https://ukdataservice.ac.uk/). NSHD data are available on request to the NSHD Data Sharing Committee. Interested researchers can apply to access the NSHD data via a standard application procedure. Data requests should be submitted to; further details can be found at http://www.nshd.mrc.ac.uk/data.aspx

https://beta.ukdataservice.ac.uk/datacatalogue/series/series?id=2000032

http://www.nshd.mrc.ac.uk/data.aspx

https://beta.ukdataservice.ac.uk/datacatalogue/series/series?id=200001

https://beta.ukdataservice.ac.uk/datacatalogue/series/series?id=2000030

https://beta.ukdataservice.ac.uk/datacatalogue/series/series?id=2000031

