## Supplementary Material for "How to mitigate selection bias in COVID-19 surveys: evidence from five national cohorts"

### Methods S1: Sensitivity analyses for social class.

We conducted sensitivity analyses for Next Steps and MCS which included both the original non-response weights as described in the COVID-19 Survey User Guide, as well as the non-response weights which we created specifically for the use of demonstrating how sample representativeness can be restored for parental social class. The original non-response weights for Next Steps and MCS included the exact same variable for parental social class which was used for the demonstration and thus had an unfair advantage relative to the multiple imputation models. However, as we can see in Fig. S5, the estimates based on the original non-response weights (blue bars) and the newly created non-response weights (orange bars) did not differ substantially. These were the newly created non-response weights used in the main paper.

### Methods S2: Sensitivity analyses for childhood cognitive ability.

We conducted sensitivity analyses for NSHD, NCDS and MCS which included both the original non-response weights as described in the COVID-19 Survey User Guide, as well as the non-response weights which we created specifically for the use of demonstrating how sample representativeness can be restored for childhood cognitive ability. The original non-response weights for NSHD, NCDS and MCS included the exact same variable for childhood cognitive ability which was used for the demonstration and thus had an unfair advantage relative to the multiple imputation models. As we can see in Fig. S6, the estimates based on the original non-response weights (blue bars) and the newly created non-response weights (orange bars) did differ, showing that estimates are not as good when no measure of childhood cognitive ability was included in the derivation of weights. We furthermore created estimates based on newly created non-response weights (purple bars) which did not include the exact same childhood cognitive ability variable as in the demonstration but did include measures of childhood cognitive ability at other time points (the same measures which were included in the equivalent multiple imputation models). With the inclusion of other cognitive measures in the derivation of weights, we were again able to restore representativeness on childhood cognitive ability to a large extend. These were the newly created non-response weights (purple bars) used in the main paper.

Similarly, we ran sensitivity analyses which included estimates based on the original multiple imputation models (which included additional measures of childhood cognitive ability from time points other than the one used in the demonstration), as well as estimates based on multiple imputation models which only included the same variables that were used in the derivation of the original non-response weights from the COVID-19 survey user guide (no additional measures of cognitive ability from other time points). As expected, the estimates from the new multiple imputation models (pink bars) did not perform as well as the imputation models which allowed for inclusion of additional measures of cognitive ability from other time points.

In conclusion, when no measures of cognitive ability were included in the derivation of weights (orange bars) or multiple imputation models (pink bars), we were not equally successful in restoring sample representativeness for childhood cognitive ability as compared to when they were included (purple and green bars). For the results in the main paper, we use non-response weights and multiple imputation models that do include these additional measures, although not the exact same childhood cognitive ability variables as were used in the demonstration itself (to avoid an unfair advantage).

### Table S1: Coding of all variables used for the derivation of weights and in the multiple imputation models.

| NSHD | NCDS | BCS | Next Steps | MCS |
|---|---|---|---|---|
| **Sex** | | | | |
| Birth<br>0 = Male / 1 = Female | Birth<br>0 = Male / 1 = Female | Birth<br>0 = Male / 1 = Female | Age 14<br>0 = Male / 1 = Female | 9 months<br>0 = Male / 1 = Female |
| **Ethnicity** | | | | |
| - | - | - | Age 14<br>1 = White / 2 = Indian, Pakistani, Bangladeshi / 3 = Black Caribbean, Black African / 4 = Mixed, Other | 9 months<br>Age 3<br>1 = White / 2 = Indian, Pakistani, Bangladeshi, Other Asian / 3 = Black Caribbean, Black African, Other Black / 4 = Mixed, Other |
| **Parental social class** | | | | |
| Age 4<br>1 = Professional, intermediate / 2 = Skilled / 3 = Partly-, unskilled<br><br>Age 11<br>1 = Professional, intermediate / 2 = Skilled / 3 = Partly-, unskilled | Birth<br>1 = Professional, managerial / 2 = Intermediate / 3 = Partly-, unskilled<br><br>Age 11<br>1 = Professional, managerial / 2 = Intermediate / 3 = Partly-, unskilled | Birth<br>1 = Professional, managerial / 2 = Intermediate / 3 = Partly-, unskilled<br><br>Age 10<br>1 = Professional, intermediate / 2 = Skilled / 3 = Partly-, unskilled | Age 14<br>1 = Managerial / 2 = Intermediate / 3 = Routine, semi-routine / 4 = Never worked | 9 months<br>1 = Managerial / 2 = Intermediate / 3 = Routine, semi-routine<br><br>Age 11<br>1 = Managerial / 2 = Intermediate / 3 = Routine, semi-routine |
| **Number of rooms at home/persons per room** | | | | |
| Birth<br>Household crowding | Birth<br>Persons per room | Birth<br>Number of rooms at accommodation | - | 9 months<br>Number of rooms at home |
| **Cognitive ability** | | | | |
| Age 8 | Age 7 | Age 10 | - | Age 5 |

| | | | | |
|---|---|---|---|---|
| Standardised score (based on Reading Comprehension, Word Reading, Vocabulary and Picture Intelligence)<br><br>Age 11<br>Standardised score (based on General Ability Test, Word Reading, Vocabulary and Arithmetic Test) | Standardised score (based on Southgate Group Reading Test, Copying Designs Test, Human Figure Drawing, Problem Arithmetic Test)<br><br>Age 11<br>Total scores of General Ability Test, Reading Comprehension Test, Mathematics Test, Copying Design Tests | Standardised score (based on British Ability Scales, Edinburgh Reading Test, Friendly Maths Test, Spelling Dictation Task, Pictorial Language Comprehension Test)<br><br>Age 5<br>Standardised score (based on English Picture Vocabulary Test, Human Figure Drawing Test, Copying Designs Test) | | Standardised score based on British Ability Scales (BAS II Naming Vocabulary, BAS II Pattern Construction, BAS II Picture Similarities)<br><br>Age 7<br>Standardised score for BAS II Word Reading<br>Standardised score for NFER Progress in Math |
| Early life mental health | | | | |
| Age 13 & 15<br>Externalising score<br>0 = absent / 1 = mild / 2 = severe<br><br>Internalising score<br>0 = absent / 1 = mild / 2 = severe | Age 16<br>Externalising symptoms standardised score;<br><br>Internalising symptoms standardised score | Age 16<br>Malaise sum score | Age 15<br>GHQ sum score | Age 11<br>SDQ Total Difficulties Score |
| Voting | | | | |
| Age 26<br>Will you vote in the next General Election?<br>0 = No / 1 = Yes | Age 42<br>Did you vote in the last General Election?<br>0 = No / 1 = Yes | Age 42<br>Voted in General Election 2010<br>0 = Didn't vote / 1 = Voted | Age 20<br>Whether voted in 2010 General Election<br>0 = Yes / 1 = No | NA |
| Membership in organisations | | | | |
| Age 43<br>0 = none / 1 = 1 / 2 = 2+ | Age 42<br>Membership in organisations<br>0 = no / 1 = yes<br><br>Membership in unions | Age 42<br>Member in organisations<br>0 = No organisations / 1 = 1 organisation / 2 = 2+ organisations | Age 26<br>Meetings for local groups/voluntary organisations<br>0 = Yes / 1 = No | Age 14<br>Youth clubs / Scouts / Etc<br>0 = At least once a month /<br>1 = Less than once a month |

| | | | | |
|---|---|---|---|---|
| | 0 = no / 1 = yes | | | |
| Internet access prior to web survey | | | | |
| Age 69<br>How often do you take part in online social networks?<br>0 = Never / 1 = Not never | Age 50<br>Whether access to internet for reasons other than work?<br>0 = yes / 1 = no | Age 46<br>Time spent on internet<br>1 = None, little / 2 = Medium / 3 = Lots | Age 26<br>Time spent on social networking website<br>0 = None / 1 = Little / 2 = Lot | Age 14<br>Use internet at home<br>1 = Little, none / 2 = Medium / 3 = Lots |
| Consent for biomarkers | | | | |
| Age 60-64<br>0 = neither, one / 1 = both | Age 44<br>0 = yes / 1 = no | Age 46<br>0 = No to one, both / 1 = Yes to both | - | - |
| Consent for linkages | | | | |
| Age 60-64<br>0 = refused / 1 = yes | - | - | Age 26<br>0 = None / 1 = Some / 2 = All | - |
| Educational qualifications | | | | |
| Age 26<br>0 = None attempted / 1 = Up to GCE 'O' Level / 2 = GCE 'A' Level / 3 = First or higher degree | Age 42<br>0 = None / 1 = NQV Level 1-3 / 2 = NVQ Level 4-5 | Age 42<br>0 = None / 1 = NQV Level 1-3 / 2 = NVQ Level 4-5 | Age 26<br>0 = None / 1 = NQV Level 1-3 / 2 = NVQ Level 4-5 | 9 months<br>0 = None / 1 = NQV Level 1-3 / 2 = NVQ Level 4-5 |
| Economic activity | | | | |
| Age 60-64<br>1 = Still in main occupation / 2 = Retired but still earning / 3 = Fully retired, Unemployed, Housewife | Age 50<br>0 = Currently employed / 1 = Not currently employed | Age 46<br>0 = Currently employed / 1 = Not currently employed | Age 26<br>0 = Currently employed / 1 = Not currently employed | Age 14<br>0 = Currently employed / 1 = Not currently employed |
| Partnership status | | | | |
| Age 69<br>1 = Single & never married / 2 = Married / 3 = Separated, divorced, widowed | Age 50<br>1 = Single & never married / 2 = Married, civil partner / 3 = Separated, divorced, widowed | Age 46<br>1 = Never married, in CP / 2 = Married, CP / 3 = Separated, divorced, widowed | Age 26<br>0 = None / 1 = Spouse, civil partner / 2 = Cohabiting partner | Age 14 (Parental partnership status)<br>1 = None / 2 = Spouse, civil partner / 3 = Separated, divorced, widowed |

| Psychological distress | | | | |
|---|---|---|---|---|
| Age 69<br>GHQ sum score | Age 50<br>Malaise sum score | Age 46<br>Malaise sum score | Age 26<br>GHQ sum score | Age 14<br>SMFQ sum score |
| BMI | | | | |
| Age 69 | Age 50 | Age 46 | Age 26 | Age 11 |
| Self-rated health | | | | |
| Age 69<br>1 = Excellent, very good / 2= Good / 3 = Fair, poor | Age 50<br>1 = Excellent, very good / 2= Good / 3 = Fair, poor | Age 46<br>1 = Excellent, very good / 2= Good / 3 = Fair, poor | Age 26<br>1 = Excellent, very good / 2= Good / 3 = Fair, poor | Age 14<br>1 = Excellent, very good / 2= Good / 3 = Fair, poor |
| Smoking status | | | | |
| Age 69<br>1 = Current Smoker / 2 = Ex-smoker / 3 = Never smoked | Age 50<br>1 = Never / 2 = Former / 3 = Current | Age 46<br>1 = Never / 2 = Former / 3 = Current | Age 26<br>1 = Never / 2 = Former / 3 = Current | Age 14<br>0 = Never smoked / 1 = Current, former, tried |
| Maternal mental health[c] | | | | |
| - | - | - | - | 9 months<br>Malaise sum score |
| Social capital/social support | | | | |
| Age 69<br>Frequency of meeting friends or relatives<br>1 = Never, almost never / 2 = Fairly frequently / 3 = Very frequently | Age 50<br>Frequency of meeting friends or relatives<br>1 = Never / 2 = Fairly frequently / 3 = Very frequently<br><br>Has people to listen to<br>1 = A little, not at all / 2 = Somewhat / 3= A great deal<br><br>Most people can be trusted<br>1 = Most people can be trusted / 2 = Can't be too careful / 3 = Other/depends | Age 46<br>Meeting family and friends<br>1 = Never, rarely / 2 = Fairly frequently / 3 = Very frequently<br><br>People around would be willing to listen<br>1 = A little, not at all / 2 = Somewhat / 3 = A great deal | Age 26<br>Meeting family and friends<br>1 = Never, rarely / 2 = Fairly frequently / 3 = Very frequently<br><br>People around would be willing to listen<br>1 = A little, not at all / 2 = Somewhat / 3 = A great deal<br><br>Trust scale (continuous, 0 - 10) | Age 14<br>Family, friends who help me feel safe, happy<br>0 = Very true / 1 = partly true, not true at all<br><br>Someone I trust<br>0 = Very true / 1 = partly true, not true at all<br><br>No one I feel close to<br>0 = Very, partly true / 1 = not true at all |

| Income | | | | |
|---|---|---|---|---|
| Age 69<br>Quantiles of total income | Age 55<br>Quantiles of total income | Age 42<br>Quantiles of total income | Age 26<br>Quantiles of total income | Age 14<br>Quantiles of total income |
| Number of non-responses across all previous sweeps | | | | |
| Birth – age 69 | Birth – age 55 | Birth – age 42 | Age 14 – age 26 | 9 months – age 14 |
| Response to COVID-19 Wave 1 survey[D] | | | | |
| Age 74 | Age 62 | Age 50 | Age 30 | Age 19 |
| Response to COVID-19 Wave 2 survey[E] | | | | |
| - | Age 62 | Age 50 | Age 30 | Age 19 |

### Fig. S1: Restoring representativeness for parental social class in COVID wave 2.

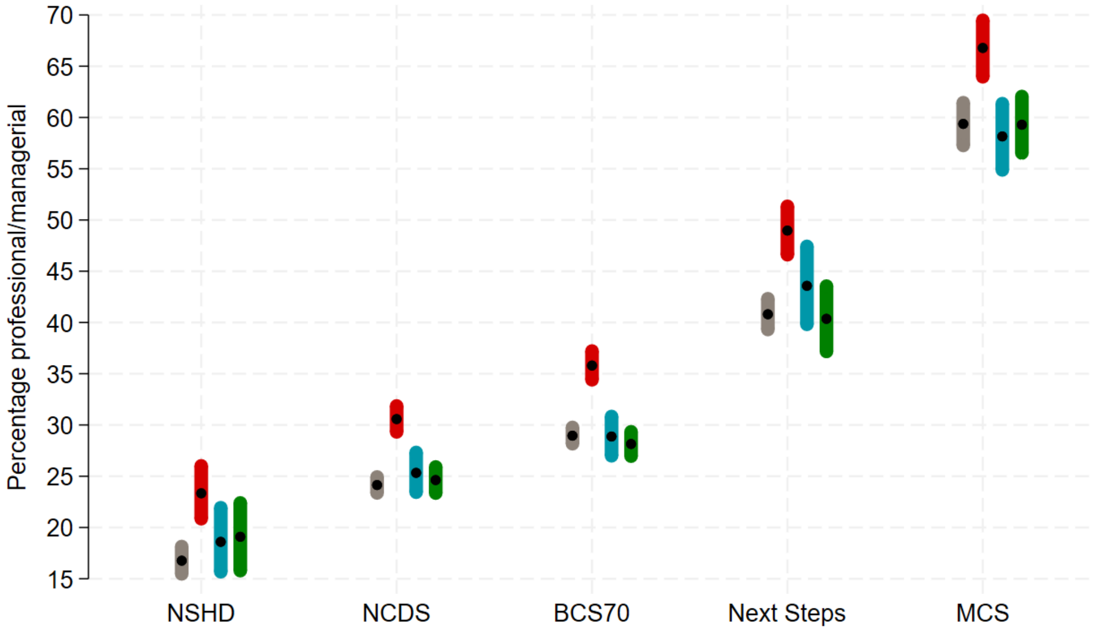

Percentage of highest social class (professional/managerial) in each cohort under different estimation approaches to account for non-response in the COVID-19 Wave 2 survey. Grey: using observed baseline data from the whole cohort; red: using observed baseline data from COVID-19 Wave 2 survey respondents only – unweighted (NCDS and BCS70) or using design weight only (NSHD, Next Steps and MCS); blue**:** using observed baseline data from COVID-19 Wave 2 survey respondents only – weighted using non-response weights (in addition to design weights as appropriate); green: using multiple imputation (plus design weights where appropriate). NSHD: National Survey of Health and Development; NCDS: 1958 National Child Development Study; BCS70: 1970 British Cohort Study; MCS: Millennium Cohort Study.

Fig. S2: Restoring representativeness for parental social class in COVID Wave 3.

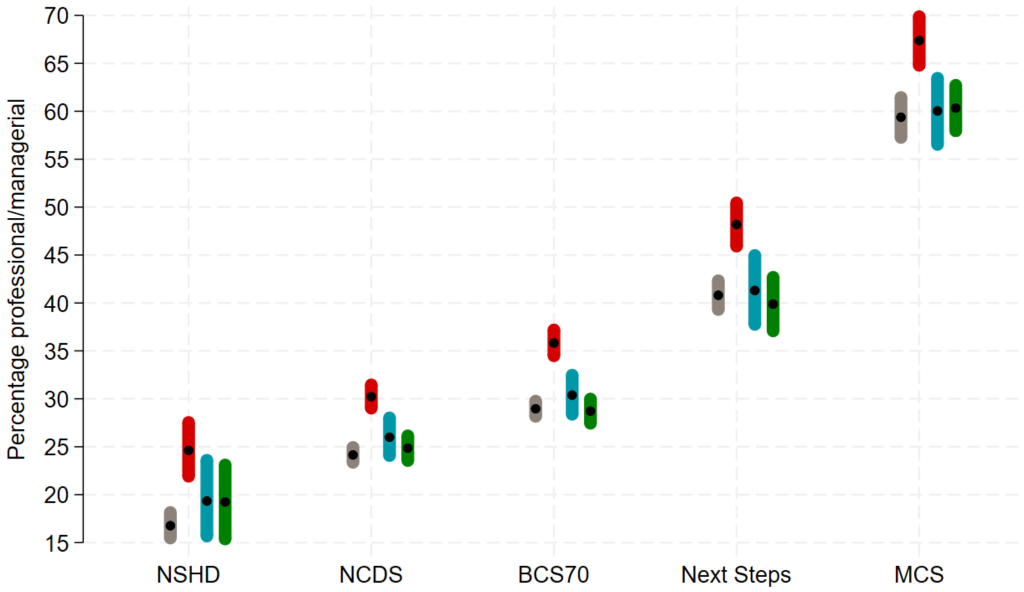

Percentage of highest social class (professional/managerial) in each cohort under different estimation approaches to account for non-response in the COVID-19 Wave 3 survey. Grey: using observed baseline data from the whole cohort; red: using observed baseline data from COVID-19 Wave 3 survey respondents only – unweighted (NCDS and BCS70) or using design weight only (NSHD, Next Steps and MCS); blue**:** using observed baseline data from COVID-19 Wave 3 survey respondents only – weighted using non-response weights (in addition to design weights as appropriate); green: using multiple imputation (plus design weights where appropriate). NSHD: National Survey of Health and Development; NCDS: 1958 National Child Development Study; BCS70: 1970 British Cohort Study; MCS: Millennium Cohort Study.

### Fig. S3: Restoring representativeness for childhood cognitive ability in COVID wave 2.

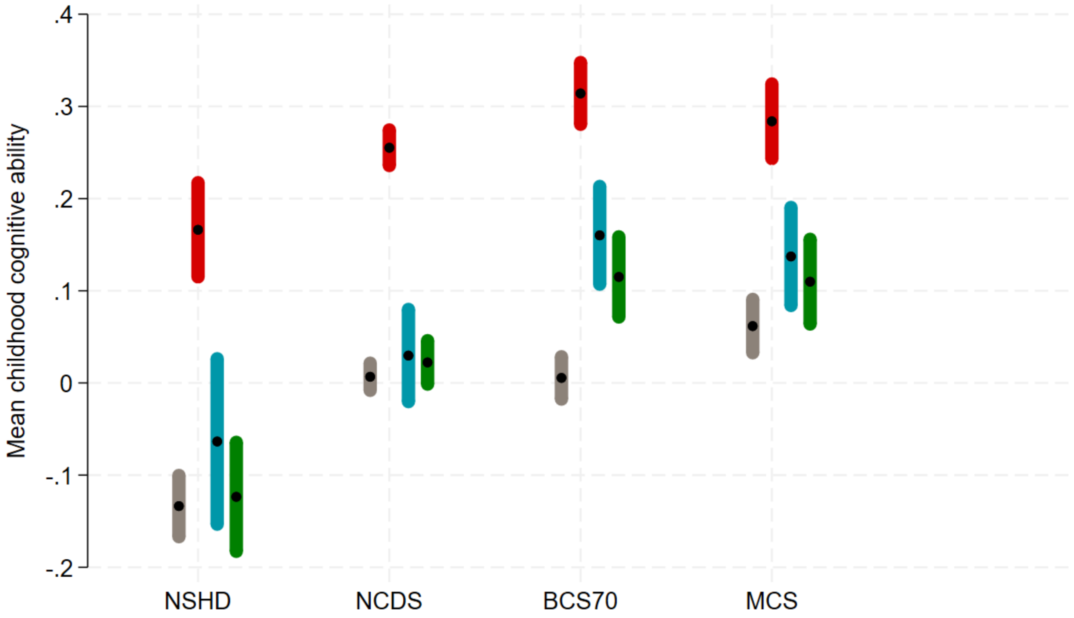

Mean of childhood cognitive ability in each cohort under different estimation approaches to account for non-response in the COVID-19 Wave 2 survey. Grey: using observed baseline data from the whole cohort; red: using observed baseline data from COVID-19 Wave 2 survey respondents only – unweighted (NCDS and BCS70) or using design weight only (NSHD, Next Steps and MCS); blue**:** using observed baseline data from COVID-19 Wave 2 survey respondents only – weighted using non-response weights; green: using multiple imputation. NSHD: National Survey of Health and Development; NCDS: 1958 National Child Development Study; BCS70: 1970 British Cohort Study; MCS: Millennium Cohort Study. Design weights were used in the estimation of means when available (NSHD, MCS) which explains why the mean of the standardised score is not always exactly 0.

### Fig. S4: Restoring representativeness for childhood cognitive ability in COVID wave 3.

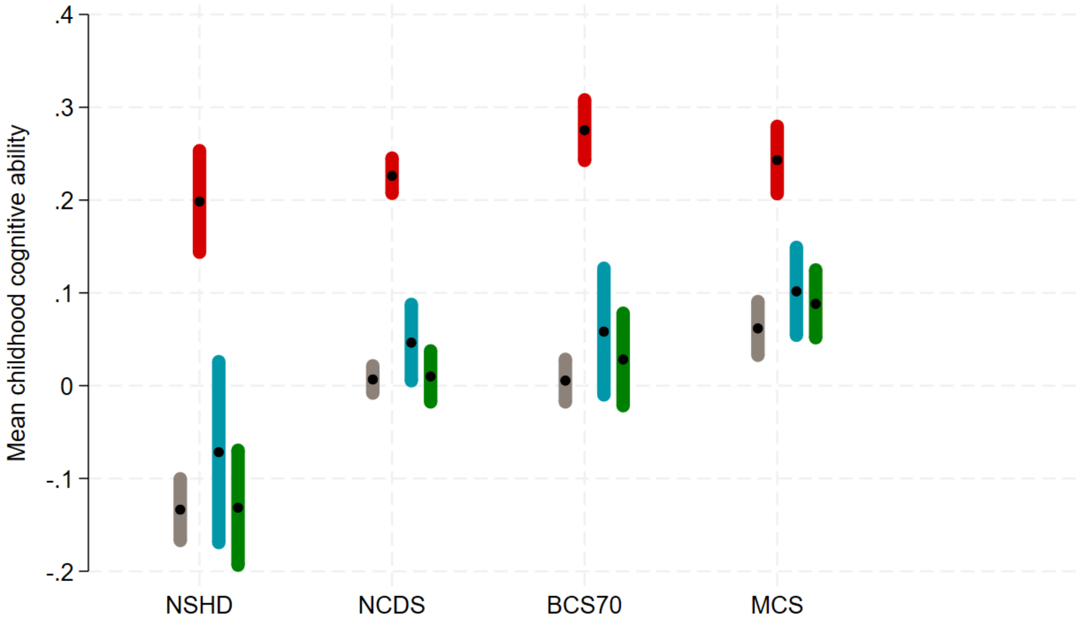

Mean of childhood cognitive ability in each cohort under different estimation approaches to account for non-response in the COVID-19 Wave 3 survey. Grey: using observed baseline data from the whole cohort; red: using observed baseline data from COVID-19 Wave 3 survey respondents only – unweighted (NCDS and BCS70) or using design weight only (NSHD, Next Steps and MCS); blue**:** using observed baseline data from COVID-19 Wave 3 survey respondents only – weighted using non-response weights; green: using multiple imputation. NSHD: National Survey of Health and Development; NCDS: 1958 National Child Development Study; BCS70: 1970 British Cohort Study; MCS: Millennium Cohort Study. Design weights were used in the estimation of means when available (NSHD, MCS) which explains why the mean of the standardised score is not always exactly 0.

### Fig. S5: Sensitivity analyses for parental social class.

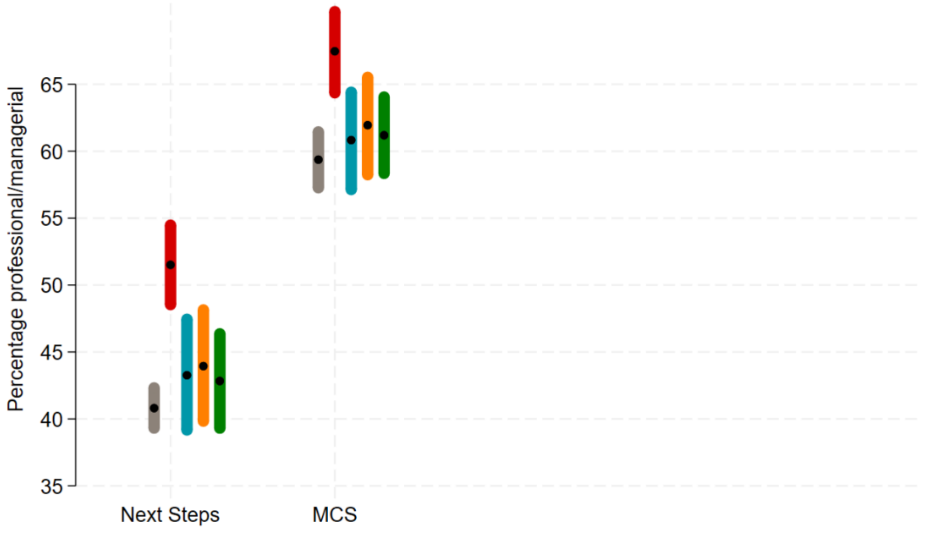

Percentage of highest social class (professional/managerial) in each cohort under different estimation approaches to account for non-response in the COVID-19 Wave 1 survey. Grey: using observed baseline data from the whole cohort; red: using observed baseline data from COVID-19 Wave 1 survey respondents only – unweighted (NCDS and BCS70) or using design weight only (NSHD, Next Steps and MCS); blue**:** using observed baseline data from COVID-19 Wave 1 survey respondents only – weighted using the original non-response weights from the COVID web survey user guide; orange: weighted using new non-response weights that do not include the same parental social class variable as in our example; green: using multiple imputation. MCS: Millennium Cohort Study.

### Fig. S6: Sensitivity analyses for childhood cognitive ability.

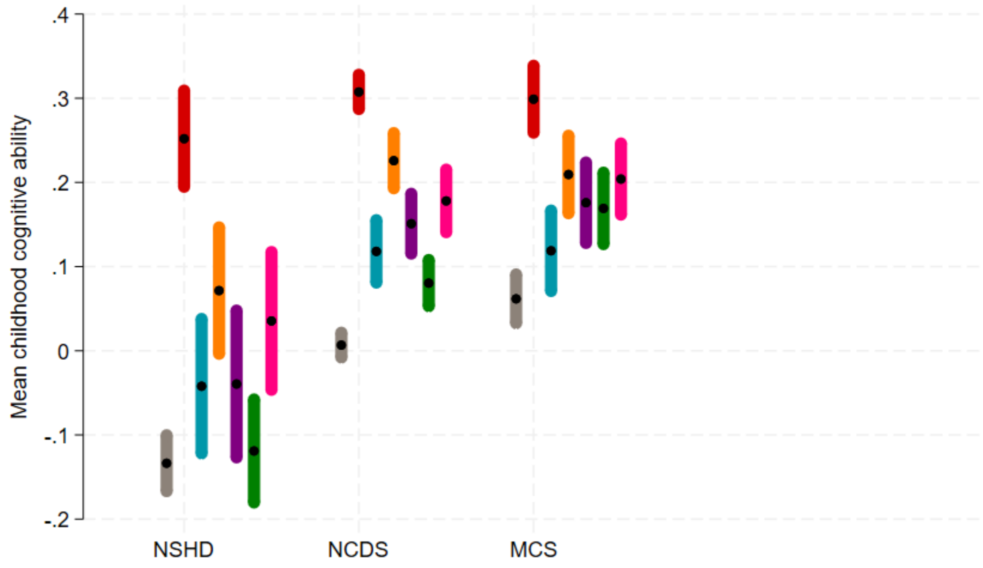

Mean of childhood cognitive ability in each cohort under different estimation approaches to account for non-response in the COVID-19 Wave 1 survey. Grey: using observed baseline data from the whole cohort; red: using observed baseline data from COVID-19 Wave 1 survey respondents only – unweighted (NCDS and BCS70) or using design weight only (NSHD, Next Steps and MCS); blue**:** using observed baseline data from COVID-19 Wave 1 survey respondents only – weighted using the original non-response weights from the COVID-19 survey user guide; orange: weighted using new non-response weights that do not include the same cognitive ability variable used in our example; purple: weighted using new non-response weights that use a cognitive ability measure from a different wave than the one in our example (same as in multiple imputation model); green: using multiple imputation; pink: using multiple imputation based on the same variables used for the derivation of the original non-response weights (which means not including cognitive ability measures from other time points). NCDS: 1958 National Child Development Study; MCS: Millennium Cohort Study. Design weights were used in the estimation of means when available (MCS) which explains why the mean of the standardised score is not always exactly 0.
